# Impact of Discharge Antihypertensive Therapy on Left-Sided Reinterventions Following the Pediatric Ross Procedure

**DOI:** 10.64898/2026.03.15.26348452

**Authors:** M. Mujeeb Zubair, Muhammad Faateh, Cameran Jones, Ayush Shah, Maria Grazia Sacco Casamassima, David Winlaw, Awais Ashfaq, Edo K. Bedzra

## Abstract

**Background:** Post-Ross procedure antihypertensive treatment strategies differ substantially, with no clear consensus and limited evidence to inform decision-making. We evaluated the association between discharge oral antihypertensive medications and post-discharge outcomes in pediatric Ross procedure patients.

**Methods:** Children (<18 years) undergoing the Ross procedure in the Pediatric Health Information System’s database (2004-2024) were included. Patients were divided into two groups based on discharge antihypertensives defined as the receipt of oral antihypertensive medication during the final two days of hospitalization: Anti-hypertensive (Anti-HTN) versus no-Anti-HTN groups. Primary outcomes were composite of left-sided (neo aortic valve/root/arch) reintervention or mortality at up to five years post-procedure. Trends in oral-antihypertensive therapy use post-Ross were examined.

**Results:** 2,097 children were included, of which 1,234 (59%) were discharged with an oral anti-hypertensive regimen. Of these, 253 (21%) were discharged on >1 oral anti-hypertensive drug class. Anti-HTN patients had lower rates of the composite of left-sided interventions or mortality at one (2.8% vs 6.1%), three (6.3% vs 9.8%) and five years (8.9% vs 13.9%), log-rank=0.0025). On stratification by age categories, statistically significant results were only observed in age category 1-12 years (log-rank=0.0127). Lowest reintervention/mortality rates were observed in patients receiving beta-blockers (log-rank=0.0112). Between 2006 and 2022, there was an increase in discharge prescription rates of beta-blockers and >1 anti-hypertensive drug class.

**Conclusions:** Following pediatric Ross procedure, discharge antihypertensive therapy was associated with a reduced composite risk particularly in the 1-12 year age group. These findings support prospective studies to define optimal antihypertensive strategies in Ross procedure patients.

## Introduction

Practice patterns for blood pressure management after the Ross procedure vary widely without consensus. In adults, some authors advocate protocols with intensive blood pressure management post-Ross procedure and suggest there is significant clinical benefit mediated by the positive effect of antihypertensives on neo-aortic root/ascending aorta dilation, and aortic stiffness.^1–3^ However no such evidence exists to guide management in children. Thus, we performed this study to evaluate the association between discharge prescription of oral antihypertensives and post-discharge outcomes (survival, readmission, and reintervention rates) in pediatric patients undergoing the Ross procedure. We hypothesized that left sided reintervention would be lower with postoperative antihypertensive use.

## Methods

A retrospective analysis of the Pediatric Health Information System’s (PHIS) database was conducted between January 1, 2004, and December 31, 2024. The PHIS is an administrative database which reports healthcare encounter level clinical, financial and pharmaceutical data from over 45 freestanding Children’s Hospitals across the United States. Each patient is assigned a unique identifier that allows longitudinal linkage to future inpatient or outpatient visits at the same hospital. Detailed information regarding PHIS can be accessed at www.childrenshospital.com. The Cincinnati Children’s Hospital Institutional Review Board approved this study, and the need for individual patient consent was waived as this was a retrospective analysis of de-identified data (CCHMC IRB #2018-6837; date of approval: October 29, 2018).

Children (<18 years) undergoing the Ross procedure were identified via International Classification of Diseases (ICD) procedure codes ICD-9 (3521, 3525, 3526) and ICD-10 (02RF07Z, 02RH07Z, 02RH08Z, 02RH0JZ, 02RH0KZ) with the additional requirement that both aortic and pulmonary valve replacement codes occurred on the same operative date. Patients experiencing in-hospital deaths or same-admission heart transplant, those with concomitant diagnosis of truncus arteriosus, connective tissue disorders, or those without pharmacy data were excluded from the analysis. Patients were divided into two groups by oral anti-hypertensive administration during the last two days prior to discharge: anti-hypertensive group (Anti-HTN group) and no-anti-hypertensive group (no-Anti-HTN group). We assumed that patients who received an oral antihypertensive during the last 2 days of hospital stay were discharged home with an antihypertensive drug prescription. The anti-HTN group included those who received at least one dose of an oral antihypertensive, defined as angiotensin-converting enzyme (ACE) inhibitors, angiotensin receptor blockers (ARB), beta-blockers, and calcium-channel blockers on the day of discharge or the day before. The primary outcome of interest was a composite of death or transcatheter or surgical intervention on the aortic valve, ascending aorta, root. Secondary outcomes included right-sided reinterventions (on the pulmonary valve/homograft or right ventricular outflow tract (RVOT)) as well as the individual components of the composite outcome. All outcomes were assessed from follow-up visits up to five years following the Ross procedure. Additionally, we examined trends in the use of oral antihypertensives during the study period.

### Statistical Analysis

Baseline comparisons between anti-HTN and no-anti-HTN groups were performed using chi-square, student t-test, or rank-sum tests as appropriate. Kaplan-Meier failure function estimates with log-rank tests were computed to compare primary and secondary outcomes with censoring applied at the time of last known contact for patients lost to follow-up. For the analysis of isolated left or right-sided reinterventions, deaths were censored. A multivariable Cox regression adjusting for age at Ross procedure and congenital heart disease diagnoses was used, with anti-HTN prescribed at discharge as the main exposure for survival and reinterventions. In addition, several sub-analyses were conducted for the primary outcome by age categories (<1year, 1-12 years, and 12-18 years) and by the anti-hypertensive drug class. A p-value of <0.05 was defined as statistically significant. All analyses were performed using Stata 18.0 (Stata Corp, College Station, TX).

## Results

A total of 2,097 children were included, of which 1,234 (58.9%) were discharged with an oral anti-hypertensive regimen. Detailed baseline characteristics are shown in Table 1. Among patients receiving anti-hypertensives, the distribution of drug classes was as follows: ACE-I (69.8%); ARB (2.8%); Beta-blocker (41.6%); Calcium Channel Blocker (8.0%). 20.5% were on antihypertensives from more than one unique drug class. At the time of the Ross procedure, children in the no anti-HTN group were younger than those in the anti-HTN group (median age 7.9 vs 8.6 years; p=0.009). However, there were no significant differences in aortic valve pathology (stenosis, insufficiency, or mixed) (p=0.59), history of endocarditis (anti-HTN 3.3% vs 2.7%, p=0.39), or rheumatic heart disease (anti-HTN 2.5% vs 2.8%, p=0.71). Anti-HTN patients were more likely to have a pre-operative history of renal failure (12.8% vs 7.8%, p<0.001) and had a longer post-operative length of stay (6 vs 5 days, p<0.001).

**Table 1.**
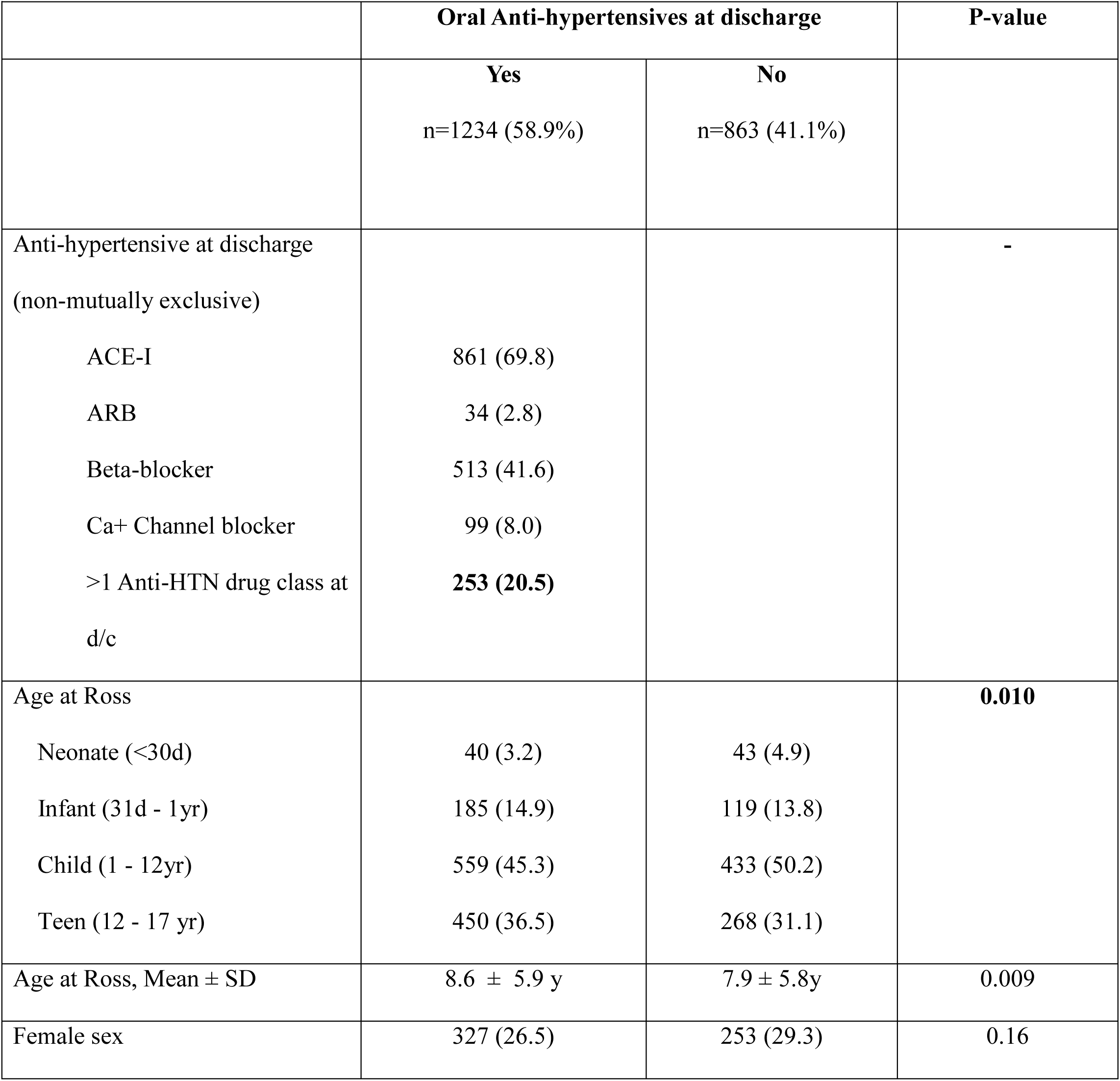
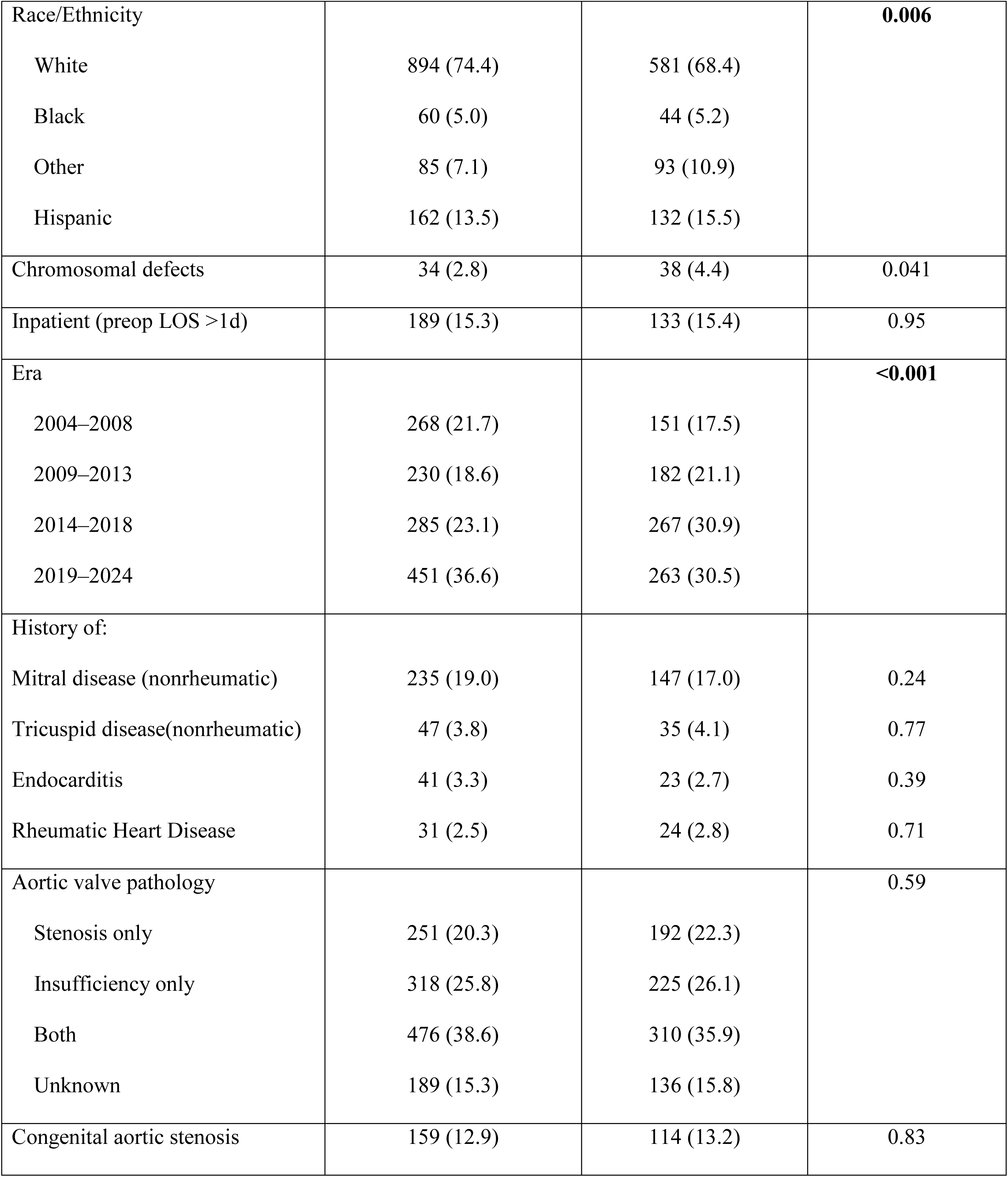
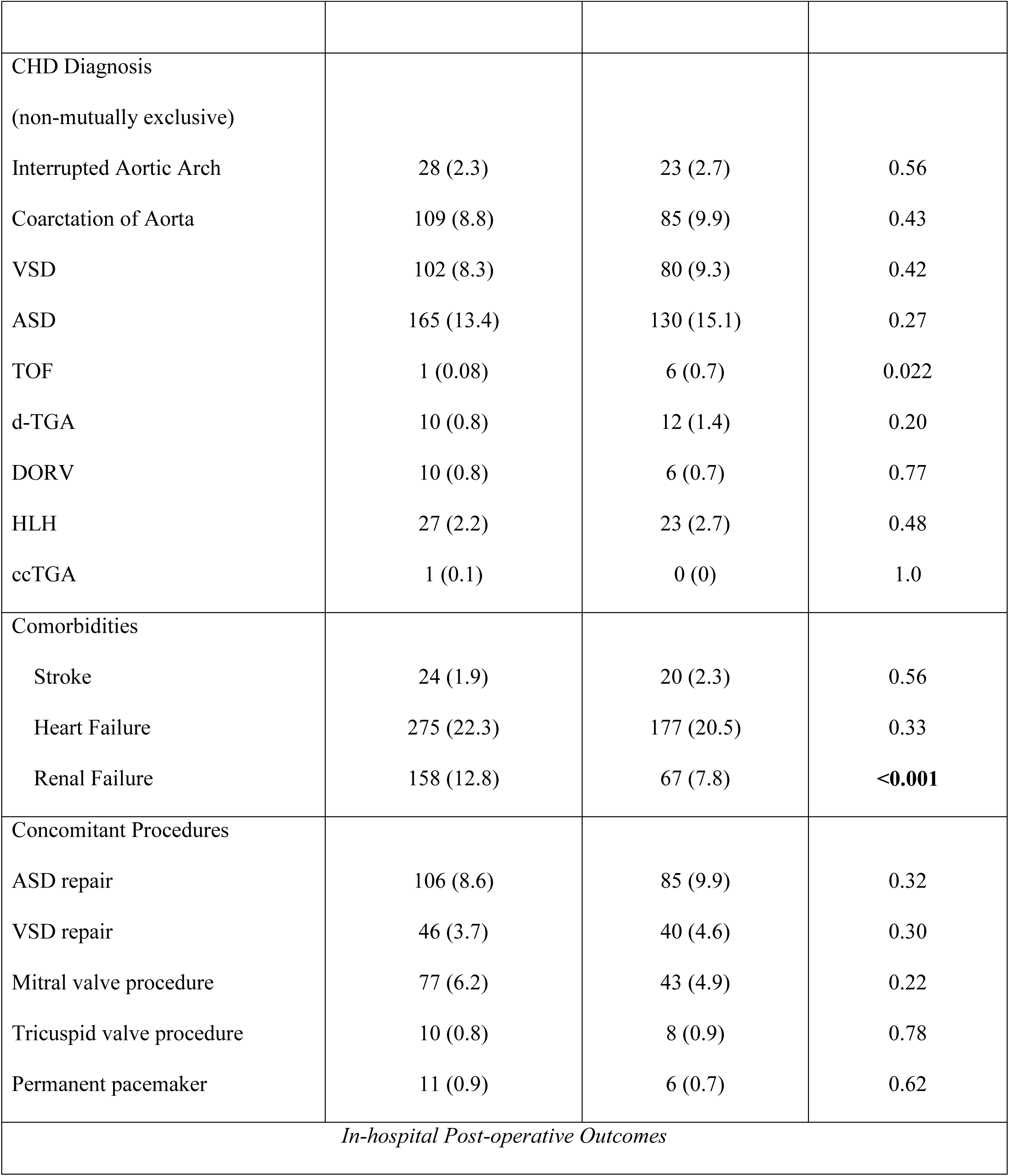
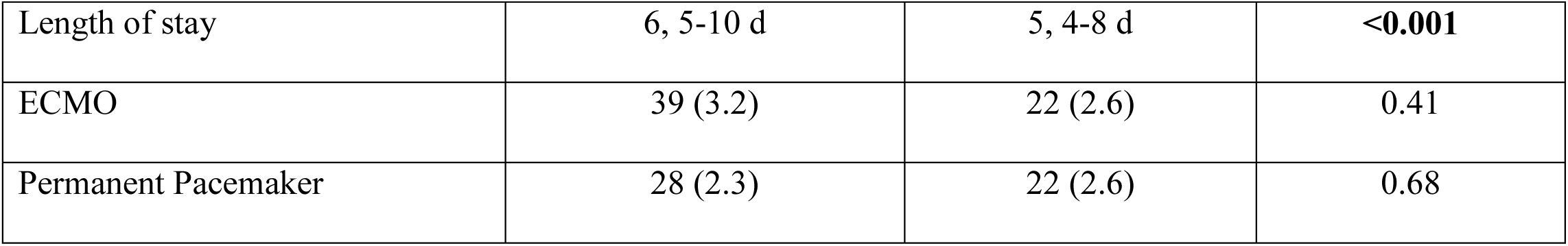
Baseline characteristics and in-hospital outcomes comparing anti-HTN and no-anti-HTN groups during index (Ross) hospitalization.

### Primary and secondary outcomes comparing anti-HTN versus no anti-HTN groups in all patients

The median follow-up was 1.4 years and was similar between the two groups. Anti-HTN patients had lower rates of the composite outcome of left-sided reintervention or death at one (2.8% vs 6.1%), three (6.3% vs 9.8%), and five year (8.9% vs 13.9%), log-rank=0.0025). Individually, the lower left-sided reintervention rate in the anti-HTN group was statistically significant (log-rank=0.0069) whereas there was no significant difference in mortality (log-rank=0.074) or right-sided reintervention rates among the anti-HTN versus no anti-HTN groups at one (2.5% vs 3.4%), three (10.8% vs 10.0%), or five (15.9% vs 18.3%) years (log-rank=0.68). (Table 2). After adjusting for age at Ross procedure and congenital heart disease diagnoses (coarctation, interrupted aortic arch, d-transposition of great arteries, tetralogy of Fallot), antihypertensive use was still associated with a lower hazard of composite of left-sided reintervention or death (HR 0.64; 95% CI, 0.48–0.84; P=0.001).

**Table 2.** Reintervention and mortality following Ross procedure comparing anti-HTN versus no-anti-HTN groups.

|  |  |  |  |
| --- | --- | --- | --- |
|  | **Anti-HTN**<br>**Group** | **No Anti-HTN**<br>**Group** | **P-value** |
| Median follow-up (yr), IQR | 1.37 (0.02-6.18) | 1.44 (0.02-6.67) | - |
| Left or reintervention or mortality composite |  |  | **0.0025** |
| 1 year (95% CI) | 2.8% (1.9–4.3) | 6.1% (4.3–8.5) |  |
| 3 years (95% CI) | 6.3% (4.7–8.9) | 9.8% (7.5–12.9) |  |
| 5 years (95% CI) | 8.9% (6.8–11.6) | 13.9% (10.9–17.7) |  |
| Left sided reintervention<br>(Neo-aortic valve, root, arch) |  |  | **0.0069** |
| 1 year (95% CI) | 2.2% (1.3-3.5) | 4.9% (3.3-7.2) |  |
| 3 years (95% CI) | 5.4% (3.8-7.4) | 8.0% (5.9-10.9) |  |
| 5 years (95% CI) | 7.7% (5.8-10.3) | 11.6% (8.9-15.2) |  |
| Post-Ross Mortality |  |  | 0.074 |
| 1 year (95% CI) | 0.6% (0.3–1.6) | 1.6% (0.8–3.2) |  |
| 3 years (95% CI) | 1.2% (0.6–2.4) | 2.3% (1.3–4.1) |  |
| 5 years (95% CI) | 1.4% (0.7–2.7) | 2.9% (1.7–5.0) |  |
| Right sided reintervention<br>(Pulmonary valve/RVOT) |  |  | 0.68 |
| 1 year (95% CI) | 2.5% (1.6-3.9) | 3.4% (2.0-5.3) |  |
| 3 years (95% CI) | 10.8% (8.5-13.5) | 10.0% (7.6-13.3) |  |
| 5 years (95% CI) | 15.9% (13.0-19.3) | 18.3% (14.8-22.7) |  |

### Primary outcome stratified by age categories and drug class

In the sub-analysis stratified by age categories, the composite outcome remained significant in patients aged between 1-12 years, comparing anti-HTN vs no anti-HTN groups at one year (1.4% vs 2.2%), three years (4.5% vs 5.4%), and five years (6.2% vs 8.9%), log-rank=0.0127. Composite outcome rates were also lower in all time points among anti-HTN patients in the neonatal/infant and >12-year age category; however, these did not reach statistical significance (>0.05). (Table 3).

**Table 3.** Sub-analysis of left-sided reinterventions or death (composite) comparing anti-HTN vs no anti-HTN stratified by age categories.

| **Age Category** | **Anti-HTN at Ross Discharge** | **1-year % (95% CI)** | **3-year % (95% CI)** | **5-year % (95% CI)** | **Log-rank** |
| --- | --- | --- | --- | --- | --- |
| **<1 year** | Yes | 3.6% (1.6–7.9) | 7.2% (4.0–12.6) | 9.8% (5.9–16.1) | 0.074 |
|  | No | 10.8% (6.4–17.9) | 15.5% (10.0–23.5) | 21.8% (14.9–31.2) |  |
| **1–12 years** | Yes | 1.4% (0.6–3.4) | 4.5% (2.7–7.5) | 6.2% (3.9–9.7) | **0.0127** |
|  | No | 2.2% (1.0–4.9) | 5.4% (3.1–9.1) | 8.9% (5.9–13.7) |  |
| **12–17 years** | Yes | 4.5% (2.5–8.3) | 8.9% (5.4–14.4) | 13.9% (8.8–21.6) | 0.26 |
|  | No | 9.6% (5.6–16.4) | 13.5% (8.5–21.2) | 17.0% (10.8–26.3) |  |

In the sub-analysis by anti-hypertensive drug category, the cumulative incidence of the composite outcome was highest in patients receiving no antihypertensive therapy and lowest in patients on beta-blocker therapy. At one, three, and five years, rates were 6.1%, 9.2%, and 13.9% for no drug, and 2.4%, 4.0%, and 6.4% for beta-blocker, respectively, with intermediate rates for ACE-I/ARB and >1 drug class. The global log-rank test comparing all groups was significant (p = 0.0112). Detailed results are shown in Table 4.

**Table 4.** Sub-analysis of left-sided reinterventions or death (composite) comparing stratified by anti-hypertensive drug class.

| **Drug Class** | **1-year % (95%**<br>**CI)** | **3-year % (95%**<br>**CI)** | **5-year % (95%**<br>**CI)** | **Log-rank** |
| --- | --- | --- | --- | --- |
| **No Drug** | 6.1% (4.3–8.5) | 9.2% (7.5–12.9) | 13.9% (10.9–17.7) | **0.0112** |
| **ACE-I or ARB** | 3.0% (1.7–5.3) | 6.4% (4.3–9.5) | 8.6% (6.0–12.2) |  |
| **Beta-blocker** | 2.4% (0.9–6.9) | 4.0% (1.8–8.9) | 6.4% (3.1–12.7) |  |
| **>1 Drug Class\*** | 3.1% (1.3–7.4) | 7.8% (4.2–14.3) | 12.3% (7.0–21.1) |  |

*Values represent Kaplan–Meier cumulative incidence estimates for left-sided reintervention or death at 1, 3, and 5 years. Single-agent CCB excluded due to small numbers; all multi-drug patients were retained in the “>1 Drug Class” category.*

### Trends in anti-hypertensive therapy prescribed at the time of pediatric Ross discharge

Overall, rates of anti-hypertensive therapy (any drug prescribed) remained relatively stable from 2006 to 2022, with 60-70% patients receiving at least one anti-hypertensive. However, significant trends were observed in the type of drug class. Rates of beta-blocker prescriptions increased from 18% to 38% at the end of the study period. Similarly, rates of multi-agent anti-hypertensive therapy increased from 6% to 28%. Rates of ACE-I prescriptions decreased from 46% to 37% during the study period.

## Discussion

In this large, multi-institutional PHIS cohort of pediatric patients undergoing the Ross procedure, discharge prescription of an oral antihypertensive was associated with a lower hazard of the composite outcome of left-sided reintervention or death at five-year follow up (adjusted HR 0.64; 95 % CI, 0.48-0.84). These findings were primarily driven by fewer left-sided reinterventions, while mortality and right-sided interventions were not different between the groups. Following the Ross procedure, the pulmonary autograft is exposed to systemic pressures, requiring adaptation to a new mechanical environment. Heart rate, aortic wall stress, and pulsatile load influence autograft sinus remodeling. Therefore, excessive stress on the implanted autograft can lead to neo-aortic root dilation and increased aortic stiffness, and new aortic insufficiency that could later require reintervention.^4–7^ Evidence from adult literature suggests that increased systemic and pulmonary artery pressure are independently associated with premature degeneration of the autograft and homograft, respectively. Therefore, strict blood pressure control has been recommended for the first 6 to 12 months postoperatively, with beta-blockade recommended as first-line therapy to reduce autograft wall stress.^6,8–10^ Conversely, there is no consensus on prophylactic antihypertensive therapy in children after the Ross procedure. Current pediatric guidelines do recommend ACE-I/ARBs as first line therapy for managing childhood hypertension.^11^ In our study, we found 60% of Ross procedure patients were on antihypertensive therapy at discharge with a trend toward decreased utilization of ACE-I from 46% to 37%, in favor of increased beta-blocker use (18% to 38%).While the mechanism for improved outcomes is unclear, this finding is welcome as patients discharged on beta-blocker therapy experienced a lower rate of the composite outcome. Perhaps the multipronged effect of beta blockade on autograft stress through reductions in heart rate, pulsatile load, and wall stress offers an advantage. Future prospective studies to guide the selection of ideal antihypertensive agent, establish the optimal dosage, and therapy duration following the pediatric Ross procedure can help shed light on these findings.

When stratified by age, children aged 1-12 years had the greatest benefit from antihypertensive therapy, as evidenced by decreased left-sided interventions. This suggests that autograft remodeling following the Ross procedure may be influenced by age.^12–15^ This potentially can explain the failure mechanism across pediatric Ross strata. Technical or anatomical factors can lead to early failure in infants and young children as they undergo more complex (e.g., Ross-Konno) operations. Growth trajectories and unaccounted for aortic pathologies may dilute the uniform effect of medications at the other end of the spectrum. Younger children, particularly infants under one, experience higher mortality and reintervention rates compared with older children.^16^ Children aged 1-12 may represent a vulnerable remodeling window, when rapid growth and ongoing autograft adaptation make systemic hemodynamic stress more consequential for root dilation and valve competence, supporting targeted prospective blood pressure studies in this age group.

This work provides the largest and only observational evidence to date linking discharge antihypertensive therapy after pediatric Ross with improved left-sided durability. The clinical implications of this study are multifaceted. First, we advocate for future prospective studies to further assess the impact of an oral-hypertensive regimen on post-Ross patients. Furthermore, it is important to delineate which antihypertensive regimen is safest and most effective in this population and to establish risk-stratified, patient-specific guidelines that factor in age, clinical status, and comorbidities to guide optimal medication selection when treatment is indicated. Our results suggest that providers should consider routine discharge prescription of antihypertensives, perhaps preferring beta-blockers in post-Ross patients between the ages of 1-12 years old.

There are some important limitations to consider which primarily stem from the retrospective analysis of an administrative database. As there is no dedicated ICD procedural code for the Ross operation, we had to infer this from a combination of existing codes. Stringent inclusion and exclusion criteria were applied to minimize the risk of misclassification. Exposure to antihypertensive therapy was defined based on medications prescribed within two days of discharge from index admission. It is important to acknowledge that antihypertensive choice is multifactorial, and the PHIS database does not capture clinical factors for such choice. The duration and intensity of antihypertensive treatment, medication adherence, changes in drug class, or crossover between treatment groups could not be captured to assess their impact on outcomes. The median follow-up duration in this study was limited and may precede the time at which neo-aortic valve-related complications following the Ross procedure typically manifest. Finally, the observed increase in length of stay (6 vs 5 days, p<0.001) among the anti-hypertensive cohort likely reflects the clinical mandate for achieving stringent blood pressure goals prior to discharge, rather than a direct correlation with surgical morbidity. Despite these limitations, this study represents a positive association between discharge antihypertensive therapy and left-sided reintervention rates following the pediatric Ross procedure and serves as an impetus for prospective studies examining the link. Additionally, medication indication could not be determined within the PHIS database. Antihypertensive agents may have been prescribed for alternative indications such as ventricular dysfunction or postoperative arrhythmias rather than hypertension alone

In conclusion, in this national PHIS cohort of pediatric patients undergoing the Ross procedure, antihypertensive therapy at discharge was associated with a significantly lower risk of left sided reinterventions or death. The association was most pronounced in children aged 1-12 years and among patients being discharged on beta-blockers. Our findings support routine use of anti-hypertensive therapy post-discharge after the pediatric Ross procedure. These findings justify future studies to define optimal therapy and duration aimed at improving the durability of the Ross procedure.

**Figure 1.**
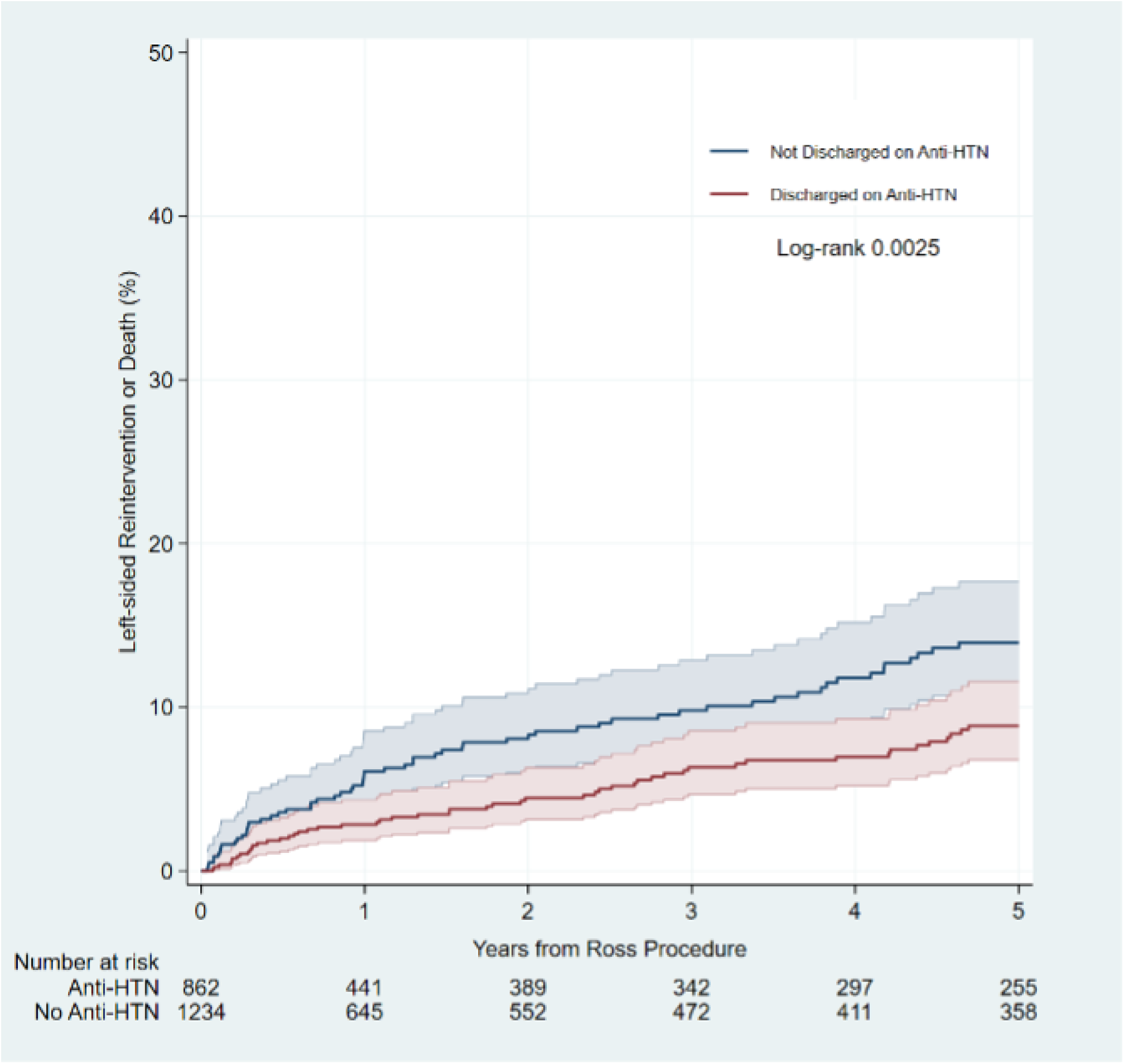
Left sided reintervention or death following pediatric Ross procedure comparing anti-HTN vs no anti-HTN groups.

**Figure 2.**
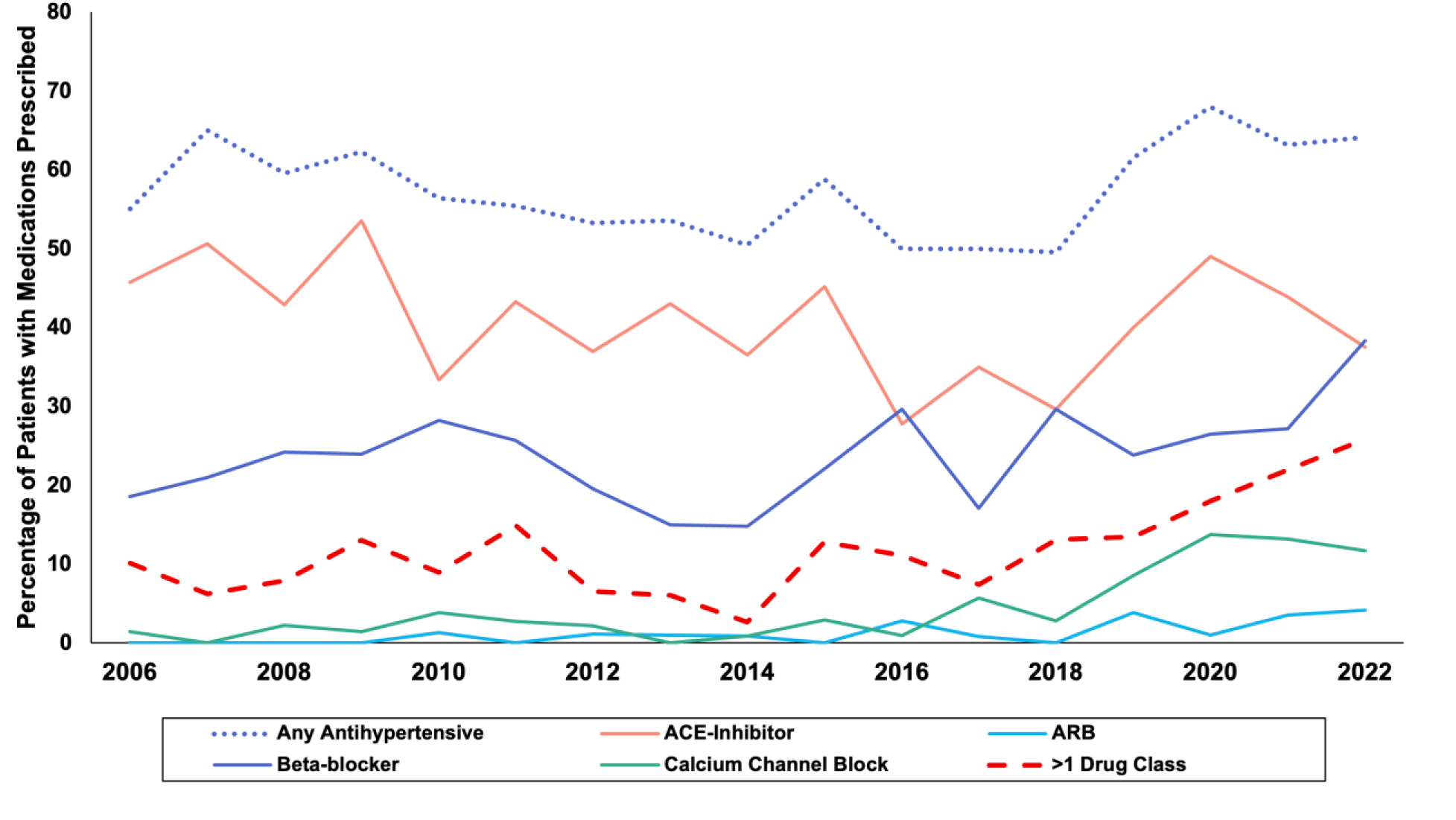
Trends in oral anti-hypertensive medications prescribed at pediatric Ross discharge.

## Data Availability

Funder requirements are not applicable, but data will be available within the article or in an appropriate repository at time of publication.

## Abbreviations

ACE: angiotensin-converting enzyme
ACE-I: angiotensin-converting enzyme inhibitor
ACC: American College of Cardiology
AHA: American Heart Association
Anti-HTN: antihypertensive
ARB: angiotensin II receptor blocker
ASD: atrial septal defect
BB: beta-blocker
CCB: calcium channel blocker
CHD: congenital heart disease
CI: confidence interval
d-TGA: d-transposition of the great arteries
DORV: double outlet right ventricle
ECMO: extracorporeal membrane oxygenation
ER: emergency room
HLH: hypoplastic left heart complex/borderline LV
HR: hazard ratio
HTN: hypertension
ICD: International Classification of Diseases
IRB: Institutional Review Board
IQR: interquartile range
LOS: length of stay
PHIS: Pediatric Health Information System
RVOT: right ventricular outflow tract
STS: Society of Thoracic Surgeons
TOF: Tetralogy of Fallot

## Acknowledgments

None

## The RECORD statement – checklist of items, extended from the STROBE statement, that should be reported in observational studies using routinely collected health data

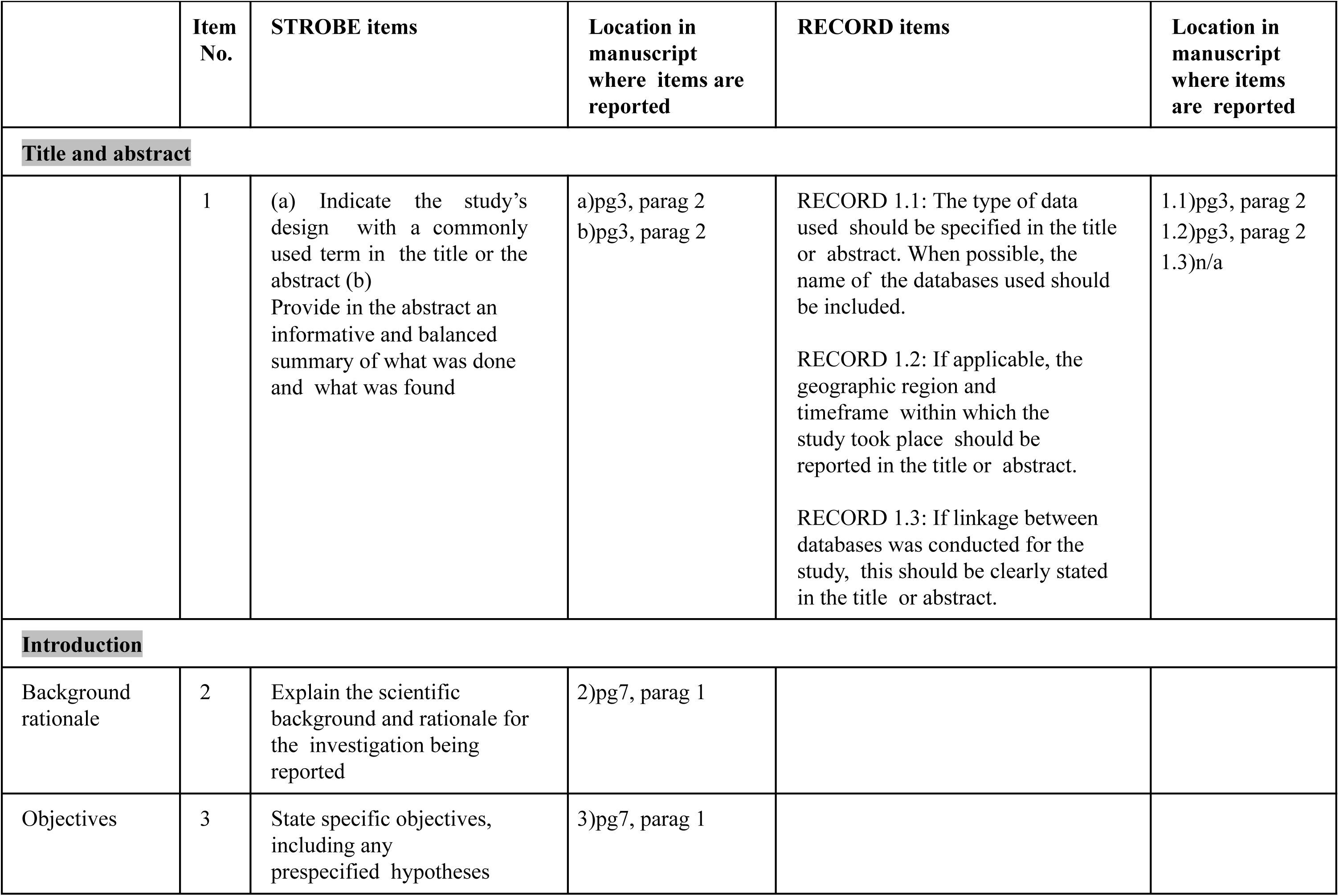
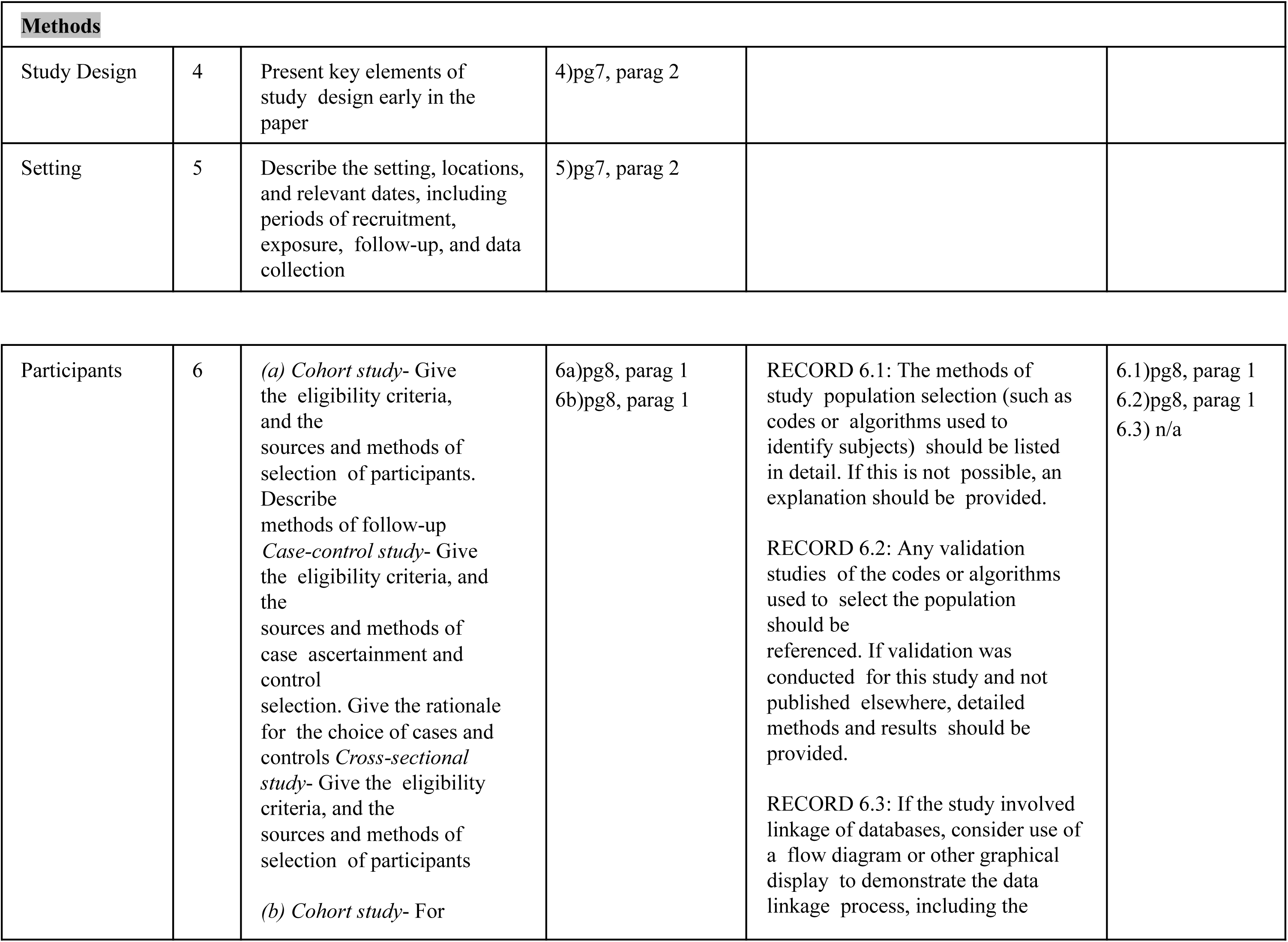
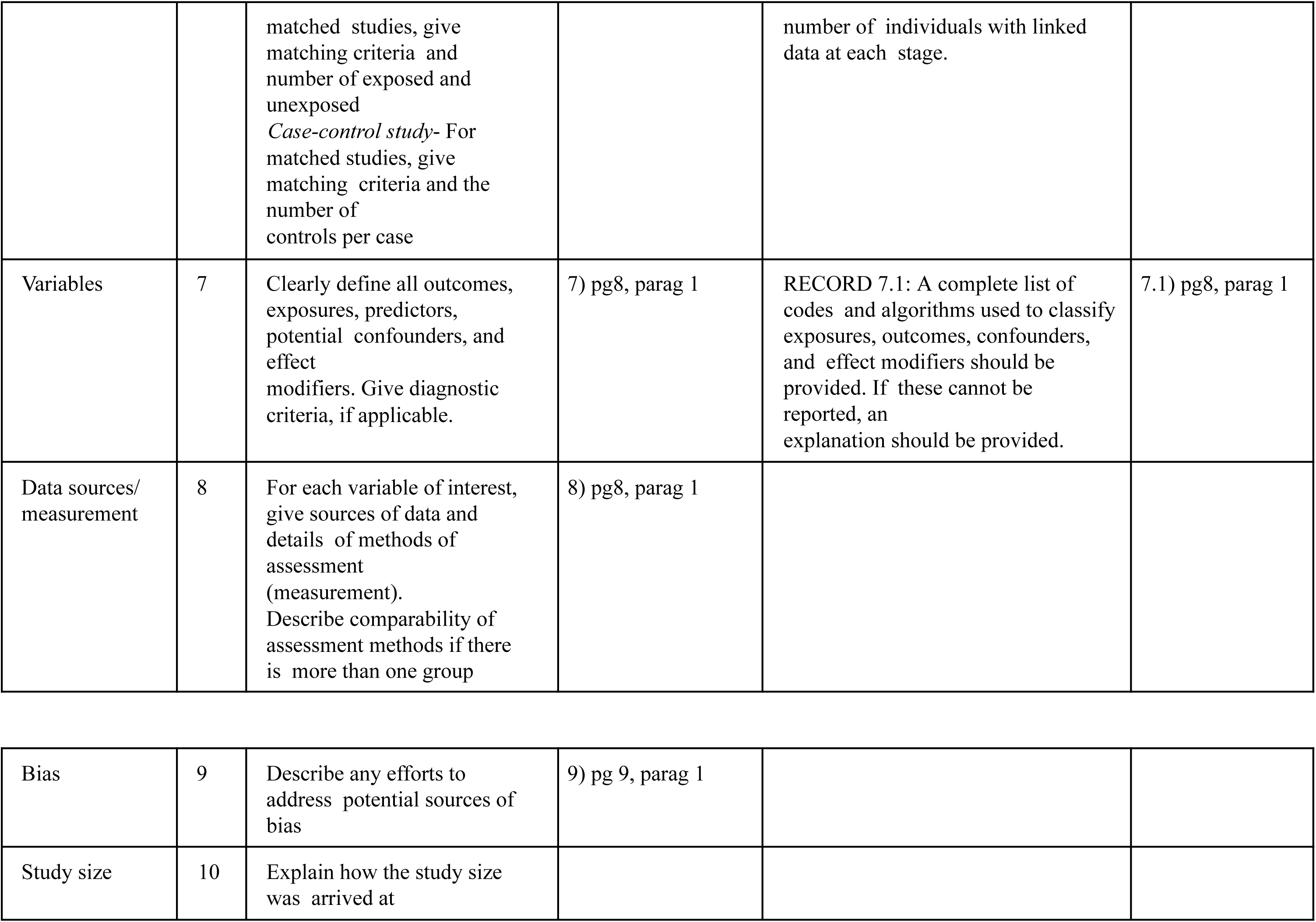
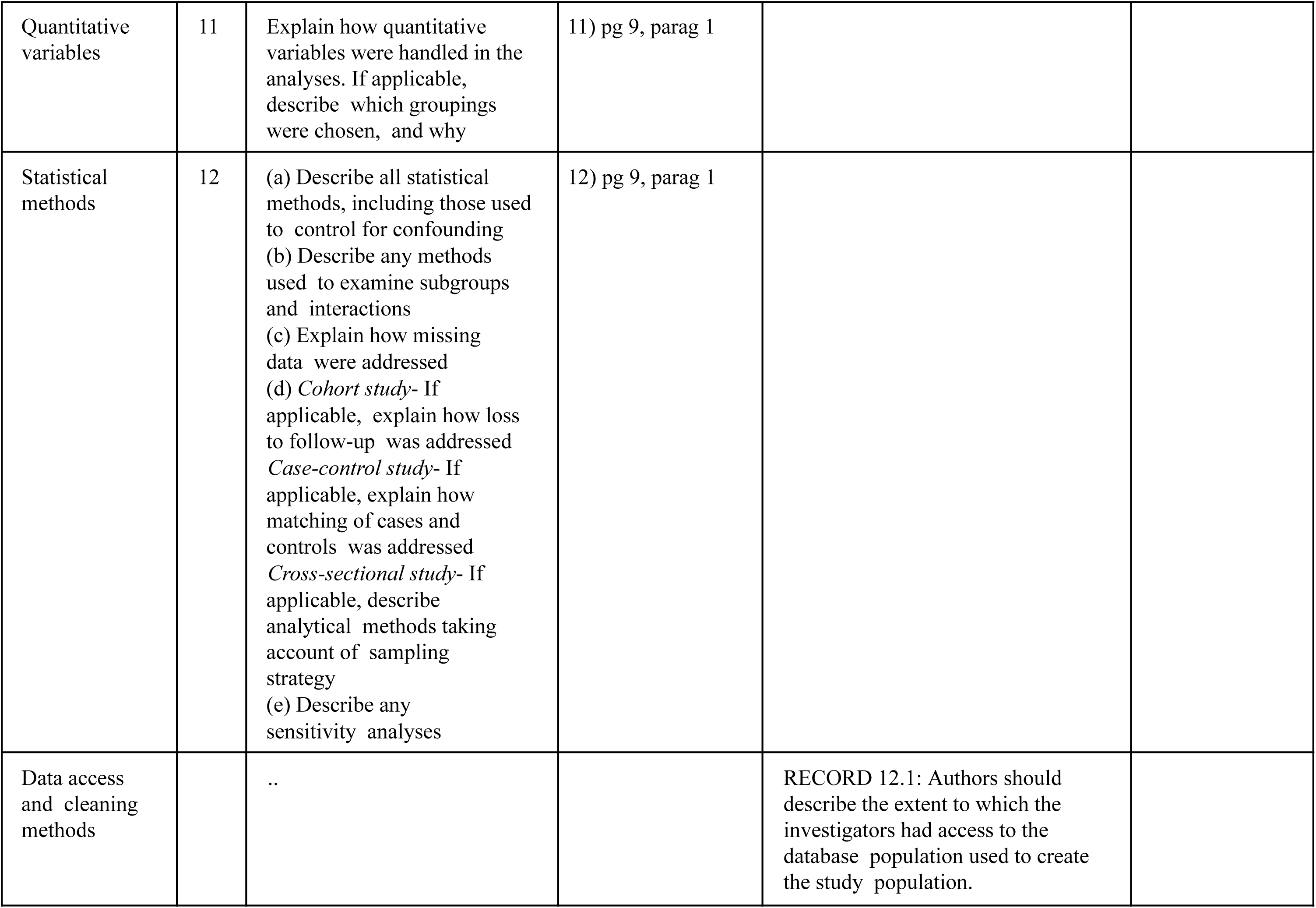
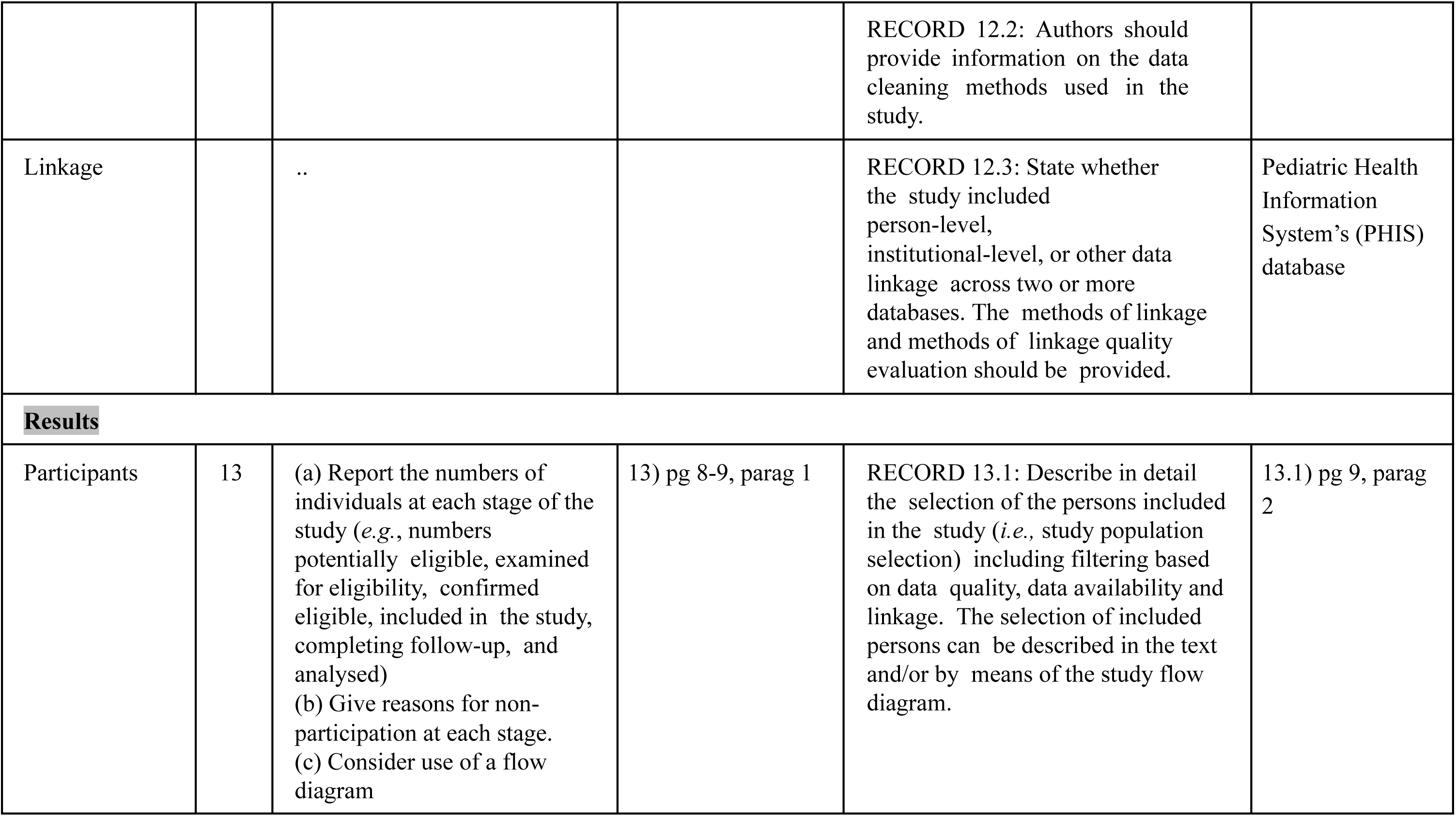
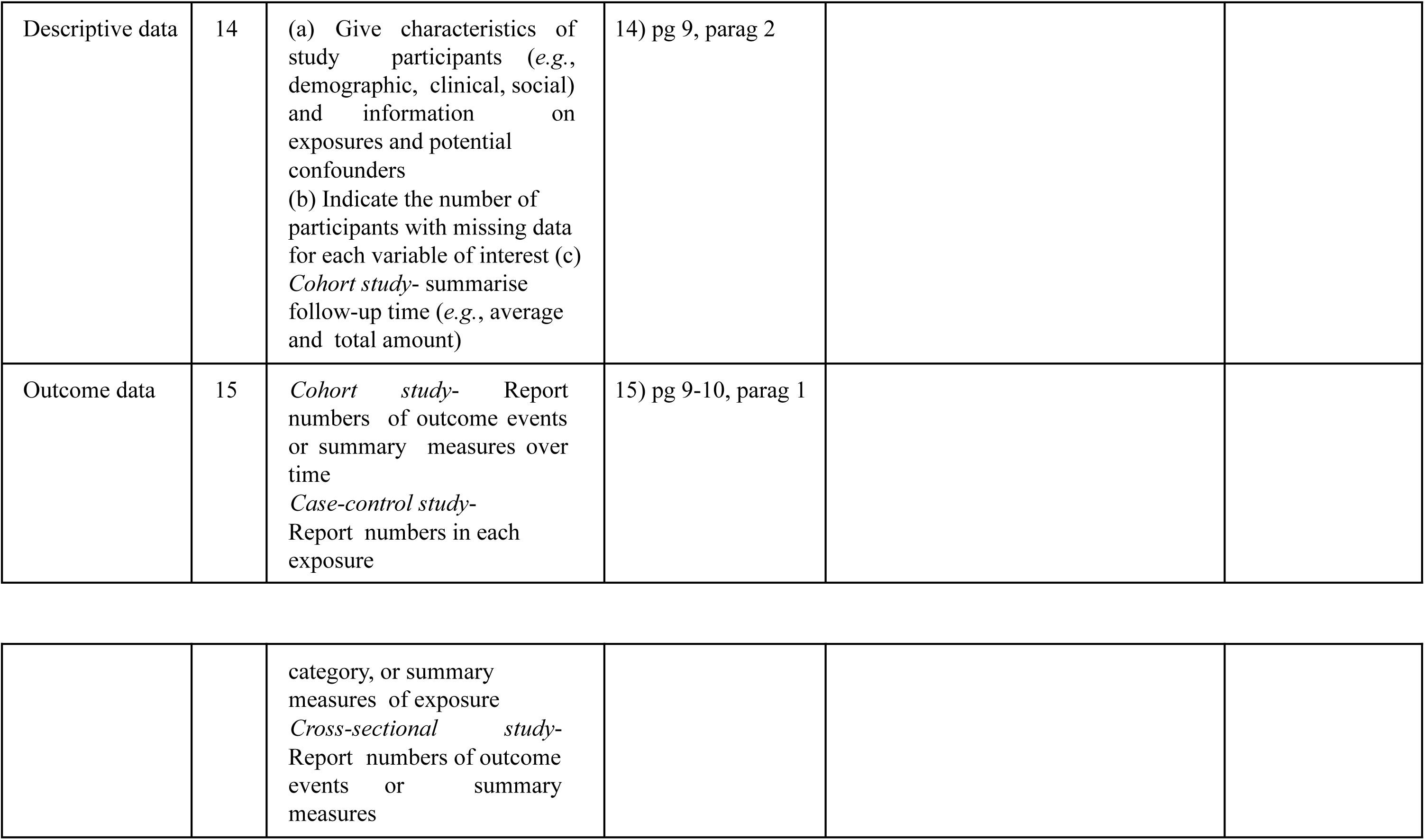
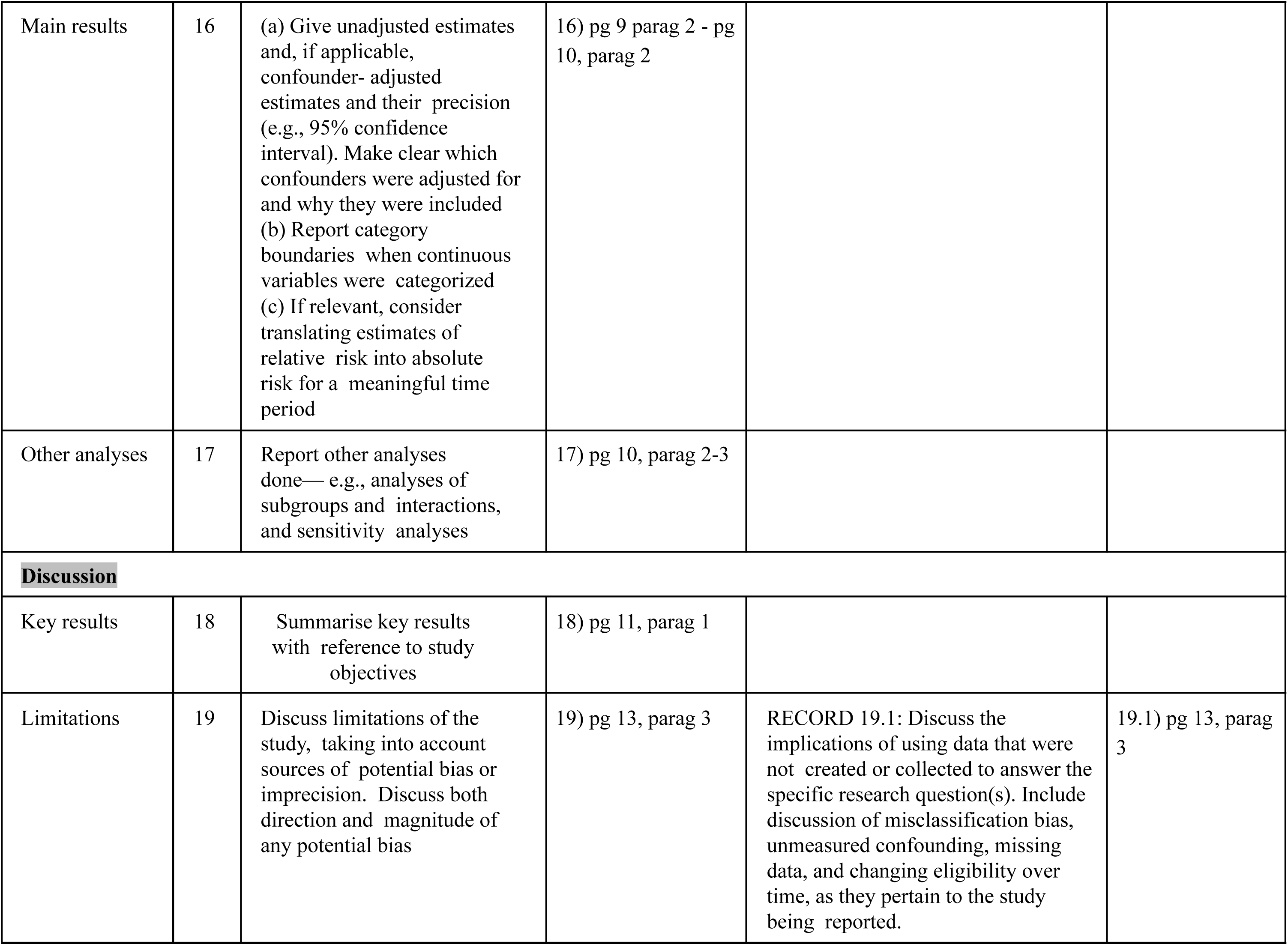
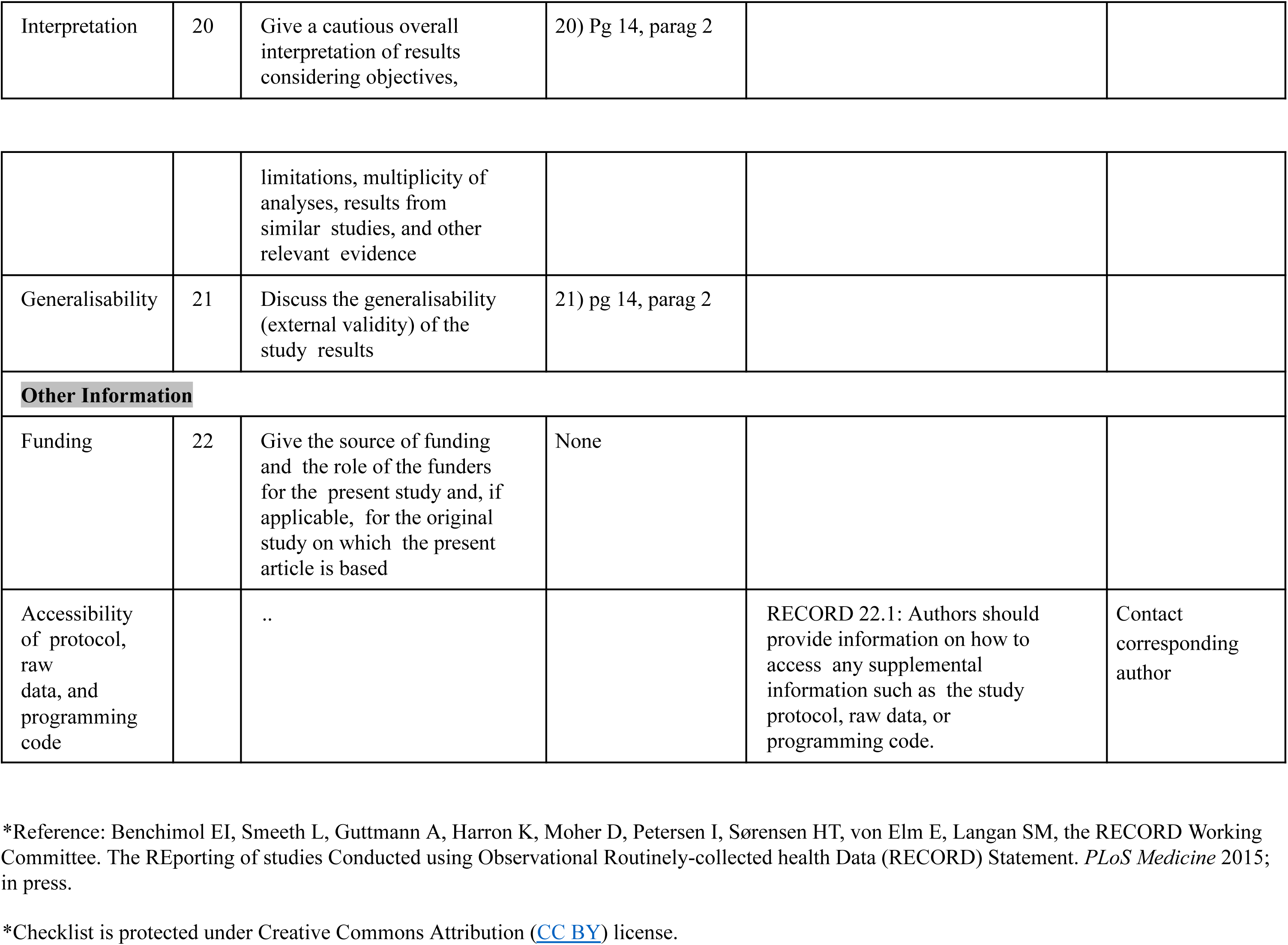

|  | Item No. | STROBE items | Location in manuscript where items are reported | RECORD items | Location in manuscript where items are reported |
| --- | --- | --- | --- | --- | --- |
| **Title and abstract** |  |  |  |  |  |
|  | 1 | (a) Indicate the study's design with a commonly used term in the title or the abstract (b) Provide in the abstract an informative and balanced summary of what was done and what was found | a)pg3, parag 2<br>b)pg3, parag 2 | RECORD 1.1: The type of data used should be specified in the title or abstract. When possible, the name of the databases used should be included.<br>RECORD 1.2: If applicable, the geographic region and timeframe within which the study took place should be reported in the title or abstract.<br>RECORD 1.3: If linkage between databases was conducted for the study, this should be clearly stated in the title or abstract. | 1.1)pg3, parag 2<br>1.2)pg3, parag 2<br>1.3)n/a |
| **Introduction** |  |  |  |  |  |
| Background rationale | 2 | Explain the scientific background and rationale for the investigation being reported | 2)pg7, parag 1 |  |  |
| Objectives | 3 | State specific objectives, including any prespecified hypotheses | 3)pg7, parag 1 |  |  |

| Methods |  |  |  |
| --- | --- | --- | --- |
| Study Design | 4 | Present key elements of study design early in the paper | 4)pg7, parag 2 |
| Setting | 5 | Describe the setting, locations, and relevant dates, including periods of recruitment, exposure, follow-up, and data collection | 5)pg7, parag 2 |
| Participants | 6 | *(a) Cohort study-* Give the eligibility criteria, and the sources and methods of selection of participants. Describe methods of follow-up<br>*Case-control study-* Give the eligibility criteria, and the sources and methods of case ascertainment and control selection. Give the rationale for the choice of cases and controls<br>*Cross-sectional study-* Give the eligibility criteria, and the sources and methods of selection of participants<br>*(b) Cohort study-* For | 6a)pg8, parag 1<br>6b)pg8, parag 1 | RECORD 6.1: The methods of study population selection (such as codes or algorithms used to identify subjects) should be listed in detail. If this is not possible, an explanation should be provided.<br>RECORD 6.2: Any validation studies of the codes or algorithms used to select the population should be referenced. If validation was conducted for this study and not published elsewhere, detailed methods and results should be provided.<br>RECORD 6.3: If the study involved linkage of databases, consider use of a flow diagram or other graphical display to demonstrate the data linkage process, including the | 6.1)pg8, parag 1<br>6.2)pg8, parag 1<br>6.3) n/a |
|  |  | matched studies, give matching criteria and number of exposed and unexposed<br>*Case-control study-* For matched studies, give matching criteria and the number of controls per case |  | number of individuals with linked data at each stage. |  |
| Variables | 7 | Clearly define all outcomes, exposures, predictors, potential confounders, and effect modifiers. Give diagnostic criteria, if applicable. | 7) pg8, parag 1 | RECORD 7.1: A complete list of codes and algorithms used to classify exposures, outcomes, confounders, and effect modifiers should be provided. If these cannot be reported, an explanation should be provided. | 7.1) pg8, parag 1 |
| Data sources/<br>measurement | 8 | For each variable of interest, give sources of data and details of methods of assessment (measurement).<br>Describe comparability of assessment methods if there is more than one group | 8) pg8, parag 1 |  |  |
| Bias | 9 | Describe any efforts to address potential sources of bias | 9) pg 9, parag 1 |
| Study size | 10 | Explain how the study size was arrived at |  |

|  |  |  |  |  |
| --- | --- | --- | --- | --- |
| Quantitative variables | 11 | Explain how quantitative variables were handled in the analyses. If applicable, describe which groupings were chosen, and why | 11) pg 9, parag 1 |  |
| Statistical methods | 12 | (a) Describe all statistical methods, including those used to control for confounding<br>(b) Describe any methods used to examine subgroups and interactions<br>(c) Explain how missing data were addressed<br>(d) *Cohort study*- If applicable, explain how loss to follow-up was addressed<br>*Case-control study*- If applicable, explain how matching of cases and controls was addressed<br>*Cross-sectional study*- If applicable, describe analytical methods taking account of sampling strategy<br>(e) Describe any sensitivity analyses | 12) pg 9, parag 1 |  |
| Data access and cleaning methods |  | .. |  | RECORD 12.1: Authors should describe the extent to which the investigators had access to the database population used to create the study population. |
|  |  |  |  | RECORD 12.2: Authors should provide information on the data cleaning methods used in the study. |  |
| Linkage |  | .. |  | RECORD 12.3: State whether the study included person-level, institutional-level, or other data linkage across two or more databases. The methods of linkage and methods of linkage quality evaluation should be provided. | Pediatric Health Information System's (PHIS) database |
| **Results** |  |  |  |  |  |
| Participants | 13 | (a) Report the numbers of individuals at each stage of the study (*e.g.*, numbers potentially eligible, examined for eligibility, confirmed eligible, included in the study, completing follow-up, and analysed)<br>(b) Give reasons for non-participation at each stage.<br>(c) Consider use of a flow diagram | 13) pg 8-9, parag 1 | RECORD 13.1: Describe in detail the selection of the persons included in the study (*i.e.*, study population selection) including filtering based on data quality, data availability and linkage. The selection of included persons can be described in the text and/or by means of the study flow diagram. | 13.1) pg 9, parag 2 |
| Descriptive data | 14 | (a) Give characteristics of study participants (*e.g.*, demographic, clinical, social) and information on exposures and potential confounders<br>(b) Indicate the number of participants with missing data for each variable of interest (c) *Cohort study*- summarise follow-up time (*e.g.*, average and total amount) | 14) pg 9, parag 2 |
| Outcome data | 15 | *Cohort study* - Report numbers of outcome events or summary measures over time<br>*Case-control study* - Report numbers in each exposure category, or summary measures of exposure<br>*Cross-sectional study* - Report numbers of outcome events or summary measures | 15) pg 9-10, parag 1 |
| Main results | 16 | (a) Give unadjusted estimates and, if applicable, confounder- adjusted estimates and their precision (e.g., 95% confidence interval). Make clear which confounders were adjusted for and why they were included<br>(b) Report category boundaries when continuous variables were categorized<br>(c) If relevant, consider translating estimates of relative risk into absolute risk for a meaningful time period | 16) pg 9 parag 2 - pg 10, parag 2 |  |  |
| Other analyses | 17 | Report other analyses done— e.g., analyses of subgroups and interactions, and sensitivity analyses | 17) pg 10, parag 2-3 |  |  |
| **Discussion** |  |  |  |  |  |
| Key results | 18 | Summarise key results with reference to study objectives | 18) pg 11, parag 1 |  |  |
| Limitations | 19 | Discuss limitations of the study, taking into account sources of potential bias or imprecision. Discuss both direction and magnitude of any potential bias | 19) pg 13, parag 3 | RECORD 19.1: Discuss the implications of using data that were not created or collected to answer the specific research question(s). Include discussion of misclassification bias, unmeasured confounding, missing data, and changing eligibility over time, as they pertain to the study being reported. | 19.1) pg 13, parag 3 |
| Interpretation | 20 | Give a cautious overall interpretation of results considering objectives, | 20) Pg 14, parag 2 |  |  |
|  |  | limitations, multiplicity of analyses, results from similar studies, and other relevant evidence |  |  |  |
| Generalisability | 21 | Discuss the generalisability (external validity) of the study results | 21) pg 14, parag 2 |  |  |
| **Other Information** |  |  |  |  |  |
| Funding | 22 | Give the source of funding and the role of the funders for the present study and, if applicable, for the original study on which the present article is based | None |  |  |
| Accessibility of protocol, raw data, and programming code |  | .. |  | RECORD 22.1: Authors should provide information on how to access any supplemental information such as the study protocol, raw data, or programming code. | Contact corresponding author |

\*Reference: Benchimol EI, Smeeth L, Guttman A, Harron K, Moher D, Petersen I, Sørensen HT, von Elm E, Langan SM, the RECORD Working Committee. The REporting of studies Conducted using Observational Routinely-collected health Data (RECORD) Statement. *PLoS Medicine* 2015; in press.
\*Checklist is protected under Creative Commons Attribution (CC BY) license.

